# Detecting Rare Diseases in Electronic Health Records Using Machine Learning and Knowledge Engineering: Case Study of Acute Hepatic Porphyria

**DOI:** 10.1101/2020.04.09.20052449

**Authors:** Aaron Cohen, Steven Chamberlin, Thomas Deloughery, Michelle Nguyen, Steven Bedrick, Stephen Meninger, John J. Ko, Jigar Amin, Alex Wei, William Hersh

## Abstract

**Background:** With the growing adoption of the electronic health record (EHR) worldwide over the last decade, new opportunities exist for leveraging EHR data for detection of rare diseases. Rare diseases are often not diagnosed or delayed in diagnosis by clinicians who encounter them infrequently. One such rare disease that may be amenable to EHR-based detection is acute hepatic porphyria (AHP). AHP consists of a family of rare, metabolic diseases characterized by potentially life-threatening acute attacks and, for some patients, chronic debilitating symptoms that negatively impact daily functioning and quality of life. The goal of this study was to apply machine learning and knowledge engineering to a large extract of EHR data to determine whether they could be effective in identifying patients not previously tested for AHP who should receive a proper diagnostic workup for AHP.

**Methods and Findings:** We used an extract of the complete EHR data of 200,000 patients from an academic medical center for up to 10 years longitudinally and enriched it with records from an additional 5,571 patients from the center containing any mention of porphyria in notes, laboratory tests, diagnosis codes, and other parts of the record. After manually reviewing all patients with the ICD-10-CM code E80.21 (Acute intermittent [hepatic] porphyria), we identified 30 patients who were positive cases for our machine learning models, with the rest of the patients used as negative cases. We parsed the record into features, which were scored by frequency of appearance and labeled by the EHR source document. We then carried out a univariate feature analysis, manually choosing features not directly tied to provider attributes or suspicion of the patient having AHP. We next trained on the full dataset, with the best cross-validation performance coming from support vector machine (SVM) algorithm using a radial basis function (RBF) kernel. The trained model was applied back to the full data set and patients were ranked by margin distance. The top 100 ranked negative cases were manually reviewed for symptom complexes similar to AHP, finding four patients where AHP diagnostic testing was likely indicated and 18 patients where AHP diagnostic testing was possibly indicated. From the top 100 ranked cases of patients with mention of porphyria in their record, we identified four patients for whom AHP diagnostic testing was possibly indicated and had not been previously performed. Based solely on the reported prevalence of AHP, we would have expected only 0.002 cases out of the 200 patients manually reviewed.

**Conclusions:** The application of machine learning and knowledge engineering to EHR data may facilitate the diagnosis of rare diseases such as AHP. The only manual modifications to this work were the removal of disease-specific or medical center specific features that might undermine our ability to find new cases. Further work will recommend clinical investigation to identified patients’ clinicians, evaluate more patients, assess additional feature selection and machine learning algorithms, and apply this methodology to other rare diseases.

## Introduction

The growing adoption of the electronic health record (EHR) worldwide has created new opportunities for leveraging EHR data for other, so called *secondary* purposes, such as clinical and translational research, quality measurement and improvement, patient cohort identification and more [1]. One emerging use case for leveraging of EHR data is to detect undiagnosed rare diseases. Although there is no absolute definition of a rare disease, the US Rare Diseases Act of 2002 defines rare diseases as those that occur in fewer than 200,000 patients worldwide [2], and the National Organization for Rare Disorders (NORD, https://rarediseases.org/) registry lists more than 1,200 diseases. Others have noted that the true number of rare diseases is unknown, and have called for more research to define them [3].

Rare diseases can be difficult to diagnose because their infrequent occurrence may result in primary care physicians not considering them in diagnostic workups. They also often have general presentations with diffuse symptoms, as well as genetic components which may require specialized testing. This lack of timely diagnosis may lead to both physical and emotional suffering as patients remain undiagnosed for prolonged periods. Additionally, a lack of accurate diagnoses increases economic burden to healthcare systems as patients continue to receive inadequate and/or inappropriate treatment. Some informatics researchers have used EHR data to detect rare diseases, such as cardiac amyloidosis [4], lipodystrophy [5], and a large collection of different diseases [6, 7].

One rare disease that may be amenable to EHR-based detection is acute hepatic porphyria (AHP). AHP is a subset of porphyria that refers to a family of rare, metabolic diseases characterized by potentially life-threatening acute attacks and, for some patients, chronic debilitating symptoms that negatively impact daily functioning and quality of life [8-12]. During attacks, patients typically present with multiple signs and symptoms due to dysfunction across the autonomic, central, and peripheral nervous systems. The prevalence of diagnosed symptomatic AHP patients is ∼1 per 100,000 [13]. Due to the nonspecific symptoms and the rare nature of the disease, AHP is often initially overlooked or misdiagnosed. A U.S. study demonstrated that diagnosis of AHP is delayed on average by up to 15 years [14].

AHP is predominantly caused by a genetic mutation leading to a partial deficiency in the activity of one of the eight enzymes responsible for heme synthesis [11]. These defects predispose patients to the accumulation of neurotoxic heme intermediates aminolevulinic acid (ALA) and porphobilinogen (PBG) when the rate limiting enzyme of the heme synthesis pathway, aminolevulinic acid synthase 1 (ALAS1), is induced [9, 15]. Gene mutations causing the disease are mostly autosomal dominant, however the disease has low penetrance (∼1%) and many specific mutations have not been identified [16]. Furthermore, families carrying the gene may have few or only one affected member. Therefore, family history can be a poor diagnostic tool for this disease. The preferred diagnostic procedure for AHP is biochemical testing of random/spot urine for ALA, PBG, and porphyrins [17, 18].

Historically, treatment of AHP has predominantly focused on avoidance of attack triggers, management of pain and other chronic symptoms, and treatment of acute attacks through the use of Panhematin^®^ (hemin for injection). Panhematin was FDA approved in 1983 for the amelioration of recurrent attacks of acute intermittent porphyria (AIP) temporally related to the menstrual cycle in susceptible women after initial carbohydrate therapy is known or suspected to be inadequate [19].

Recently, a new drug Givlaari^®^ (givosiran), for subcutaneous injection has been approved by the FDA for the treatment of adults with AHP. Givosiran is a double-stranded small interfering RNA (siRNA) molecule that reduces induced levels of the protein ALAS1. A Phase 1 trial has been published [20] and a Phase 3 randomized control trial has shown this therapy to be effective in reducing the occurrence of acute attacks and impacting other manifestations of the disease [21].

Oregon Health & Science University (OHSU) is the only academic medical center in Oregon and is thus a referral center for rare diseases like AHP. The OHSU Research Data Warehouse (RDW) is a research data “honest broker” service that provides EHR data to researchers, with appropriate IRB approval. The investigators have an ongoing institutional review board (IRB) approval to use an extract from the Oregon Health & Science University (OHSU) EHR research data warehouse (RDW) for a series of patient cohort identification projects. For this research, the patient cohort to identify was defined as those patients who have a documented clinical history of AHP, or a clinical history indicating that AHP diagnostic testing may be appropriate. The goal of this study was to apply machine learning and knowledge engineering to a large extract of EHR data to determine whether the combined approach could be effective in identifying patients not previously tested for AHP who should receive a proper diagnostic workup for AHP. This study protocol was approved by the OHSU Institutional Review Board (IRB00011159).

## Materials and Methods

### Dataset

A large dataset of approximately 200,000 patient records was requested from the RDW, complete as of the data pull date in March 2019, including over 30 million text notes plus other document types. These records corresponded to patients who had more than one primary care health care visit at our institution. Each patient record was represented as a collection of documents of types given in **Table 1**. Patient records could include zero or more documents of each type.

**Table 1.** Electronic Health Record (EHR) document types used in this research.

|  |
| --- |
| Administered Medications |
| Current Medications |
| Demographics |
| Encounter Diagnosis |
| Hospital Encounters |
| Lab Results |
| Medications Ordered |
| Microbiology Results |
| Notes |
| Problem List |
| Procedures Ordered |
| Lab Result Comments |
| Surgeries |
| Age |

To insure an adequate number of number of patients to make predictive models robust, we enriched the data set for possible AHP by adding records from an additional 5,571 patients who met one or more of the following case-insensitive criteria (see **Table 2**):

**Table 2.** Electronic Health Record (EHR) document counts of porphyria codes and mentioned in text notes or label tests.

| **Code** | **Total** | **Unique** |
| --- | --- | --- |
| ICD9 277.1 | 3879 | 308 |
| E80.0 Hereditary erythropoietic porphyria | 472 | 37 |
| E80.1 Porphyria cutanea tarda | 783 | 77 |
| E80.20 Unspecified porphyria | 2010 | 247 |
| E80.21 Acute intermittent (hepatic) porphyria | 1016 | 47 |
| E80.29 Other porphyria | 109 | 24 |
| E80.4 Gilbert syndrome | 3197 | 366 |
| E80.6 Other disorders of bilirubin metabolism | 9502 | 2308 |
| E80.7 Disorder of bilirubin metabolism, unspecified | 75 | 58 |
| Patients with porphyria mentioned in a lab test: | 359 | 175 |
| Searching field NOTE TEXT for term porphyria: | 14353 | 3012 |

- Diagnosis including “porph” in the diagnosis name
- Medication including “hemin” in the medication name
- Procedure including “porph” in the procedure name
- Clinical or result note including “porph” in the note text

To develop a gold standard for the data, a medical student (MN), overseen by clinical experts among the rest of the authors, identified patients with a high likelihood of AHP. We manually reviewed all the patients with the ICD-10-CM code E80.21 (Acute intermittent [hepatic] porphyria) in their record, looking for positive confirmation of AHP either through a lab test or a specific comment in a progress note. This process yielded 30 positive cases from the 47 coded for E80.21. As OHSU is the only academic medical center in Oregon and is thus a referral center for rare diseases like AHP, this may explain why the number of identified AHP patients in our database was higher than that which would be expected based on the global prevalence of AHP. The rest of the records were then assumed to be negative for AHP for the purposes of statistical analysis and machine learning.

We then deconstructed each patient record into a number of features to be used for machine learning. Structured data fields were encoded directly with the entire field content used as the feature. Free-text fields were parsed into unigrams and bigrams. All features were labeled with their source document. This enabled, for example, ICD-10-CM codes in the problem list to be distinguished from the same ICD-10-CM codes appearing in an encounter diagnosis. Feature values were encoded as the number of occurrences in the entire record for the patient. A summary of the types and counts of documents in the data set is shown in **Table 3**.

**Table 3.** Summary of document types and counts used in the EHR data set for this research.

| Type | Patients | Encounters | Records | Mean | Median | Max |
| --- | --- | --- | --- | --- | --- | --- |
| current_medications | 187,724 | N/A | 99,602,443 | 530.58 | 89 | 57,406 |
| demographics | 204,413 | N/A | 204,413 | 1.00 | 1 | 1 |
| encounter_attributes | 204,412 | 19,589,057 | 19,589,057 | 95.83 | 43 | 3,335 |
| encounter_diagnoses | 202,843 | 10,113,657 | 52,295,188 | 257.81 | 69 | 27,215 |
| hospital_encounters | 145,551 | 1,163,284 | 1,163,284 | 7.99 | 3 | 520 |
| lab_results | 172,795 | 2,012,185 | 58,386,934 | 337.90 | 84 | 27,384 |
| medications_ordered | 190,256 | 3,964,120 | 15,155,203 | 79.66 | 23 | 7,041 |
| microbiology_results | 54,798 | 145,528 | 1,988,429 | 36.29 | 5 | 5,174 |
| notes | 204,161 | 10,014,987 | 28,938,900 | 141.75 | 56 | 14,933 |
| problem_list | 181,221 | N/A | 1,737,749 | 9.59 | 6 | 204 |
| procedures_ordered | 198,833 | 5,129,756 | 19,501,225 | 98.08 | 31 | 35,364 |
| result_comments | 131,104 | 896,896 | 1,542,279 | 11.76 | 4 | 1,765 |
| surgeries | 44,238 | 78,403 | 83,535 | 1.89 | 1 | 54 |
| vitals | 199,971 | 3,500,418 | 18,268,032 | 91.35 | 24 | 9,442 |
| administered_medications | 100,565 | 349,332 | 17,160,858 | 170.64 | 17 | 53,178 |
| ambulatory_encounters | 204,235 | 12,091,755 | 12,091,755 | 59.21 | 27 | 1,991 |

### Machine Learning Model Feature Selection and Training

Features to be included in the machine learning model were then selected by performing univariate analysis of the entire feature set, using the confirmed AHP patients as positive samples and the rest of the data set as negative samples. For each document type, the 100 top features were chosen, ranked by odds ratio, having a p-value < 0.01 and occurring in at least 4 positive case patient records.

From these several hundred features, a manual review process was performed to ensure that none of these features were directly connected to a diagnosis of AHP, mention of AHP in the record, or treatment of AHP. This process eliminated all text features mentioning any bigram of “acute hepatic porphyria,” medications such as hematin, and laboratory codes that in the OHSU system represented tests specifically for the diagnosis of porphyria.

This process reduced the set to approximately 200 features. These features were then evaluated by using them in a machine learning model and scoring the model using 5 repetitions of 2-fold cross-validation. These experiments found that an SVM with the radial basis function (RBF) kernel scored best for the ranking metrics AUC and average precision. Linear SVM, random forests, Adaboost, J48, and several topologies of Neural Network were also tried but failed to perform as well as the RBF SVM. It was also determined that feature values were best encoded using log normalization, transforming feature occurrence counts into values between 0.0 and 1.0. Binary encoding, as well as linear normalization, failed to perform as well. We used the SVMLight implementation of the RBF kernel. Experimentation with cross-validation showed gamma = 0.04 to be optimal.

After algorithm selection, a second round of feature screening was performed. Any features with non-zero weights in the SVM model were removed if any direct connection to AHP could be established. This was performed by close scrutiny and discussion with clinical experts on each feature. For example, based on case series evidence, clinical hematology AHP specialists sometimes use cimetidine to treat AHP symptoms, as it is known to block a portion of the heme synthesis pathway as a side effect [22]. We found that cimetidine was a highly weighted feature in our initial models (due to its use by a specialist [TD] at OHSU based on case report data [22]) that had to be removed as it is given in response to AHP rather than being predictive. This process resulted in 146 total features being included in the final model.

The 146 features included in the final model are shown in **Table S-1**. Final feature set cross-validation performance on the entire training set is shown in **Table 4**.

**Table 4.** Cross-validation performance of the final feature set on the entire data set for ranking the 30 confirmed cases of porphyria higher than the general population. SVM with radial basis function (RBF) kernel and gamma = 0.04.

| Metric | Score |
| --- | --- |
| AUC | 0.775 |
| Average Precision | 0.060 |
| Precision @ 100 | 0.031 |
| Log Loss | 0.404 |

### Machine Learning for AHP Prediction and Evaluation Methodology

A final trained model using the features selected was created by training the mode on the entire data set. This model was then applied back to the entire data set in order to create an AHP prediction score for each patient. The classifier margin distance was taken as the prediction score.

The patient prediction scores were then analyzed. In particular, the range of scores obtained for the 30 confirmed positive training cases were compared to the rest of the patients in the data set. About 22,000 patients in the general population had scores that overlapped with those of the 30 positive patients. While this was only 10% of the patient records, it was more than could be manually reviewed. We decided to review the top scoring 100 cases manually from each of two subsets of the general population.

The first reviewed subset of 100 patients were those with no mention of porphyria in their chart, no related ICD-9-CM or ICD-10-CM codes, and no porphyria specific lab test. We selected the top scoring 100 patients that met these criteria. This represents the most important target population for our project – patients with persistent symptoms that have not had AHP considered and tested to rule it in or out as a diagnosis. Manual review of these cases is intended to demonstrate the potential of our proposed approach to identify potential cases of AHP that would benefit from diagnostic testing and follow up.

The second reviewed subset of 100 patients were those <u>with</u> a mention of porphyria in the text notes in their chart, but no related ICD-9-CM or ICD-10-CM diagnosis codes, and no porphyria-specific lab test. These are patients where porphyria may have been considered by the clinician, or may have been tested at another health care facility with unavailable records, or may have been a work up in progress. Manual review of these cases was intended to discern the clinical face validity of the algorithmic predictions, that is, the high scoring patients in this group score high because the algorithm is paying attention to some of the same non-AHP-specific clinical symptoms and other variables as the clinician. While the manual review of these patients was primarily intended for gaining insight into how the algorithm was scoring patients with porphyria mentioned in the charts, based on the manual review some patients who may benefit from diagnostic testing could be found.

A clinically trained reviewer assessed the patients’ records in these two non-overlapping subsets for symptom patterns consistent with acute hepatic porphyria (AHP). The reviewer was blinded to the model features. Clinical notes were searched for the ‘classic triad’ of AHP symptoms: abdominal pain, central nervous system abnormalities, and peripheral neuropathy [23]. In addition, any report of pain was assessed, and searches were also conducted for the highest incident AHP symptoms: abdominal pain, vomiting, constipation, muscle weakness, psychiatric symptoms, limb, head, neck, or chest pain, hypertension, tachycardia, convulsion, sensory loss, fever, respiratory paralysis, diarrhea [23]. All major comorbidities were also reviewed and documented, as well as alternative diagnoses to explain AHP symptom profiles.

The 100 patients with no mention of porphyria in their EHR record were classified into one of three categories: *AHP diagnostic testing likely indicated, AHP diagnostic testing possibly indicated*, and *AHP diagnostic testing unlikely indicated*. To be classified as *likely*, symptoms had to be present in all three categories of the ‘classic triad’, without a cause identified in the EHR, and with a substantial history of symptoms. To be classified as *possibly*, symptoms had to be present in at least one of the three categories, without a cause documented and with a substantial history. Patients were classified as *unlikely* if their symptoms could be explained by another diagnosis, or if they did not have a strong AHP symptom profile.

The 100 patients who did have a mention of porphyria in their clinical notes were classified into one of five categories of AHP status based on chart review and details in the clinical notes: *AHP already suspected, AHP already suspected but ruled out, diagnostic testing likely indicated but AHP not suspected, unlikely AHP*, and *AHP diagnosis mentioned in notes*. A patient was classified as *AHP already suspected* if there was any level of AHP suspicion mentioned in their clinical notes, without a formal diagnosis or lab test. *AHP already suspected but ruled out* was assigned if there was a suspicion of AHP in the note, but had been ruled out, usually by negative lab tests. These lab tests were only documented in the note, since we excluded patients from this subset who had lab tests in the laboratory data itself. *Diagnostic testing likely indicated but AHP not suspected* was assigned if there were symptoms present in at least one of the three triad categories, without a cause, but no suspicion of AHP mentioned in the notes. For these patients the clinical notes contained the string ‘porph’ but presence of ‘porph’ in the clinical note was not related to suspicion of AHP. *Unlikely AHP* was assigned if AHP type symptoms could be explained by another diagnosis, or there was not a strong AHP symptom profile. Finally, patients were assigned to *AHP diagnosis* if there was any mention of an existing AHP diagnosis in the notes, even patient reported. The reasons for the presence of the string ‘porph’ in the clinical note for the second set of 100 patients was also reviewed and documented. Patient’s categorized as *AHP already suspected* and *Diagnostic testing likely indicated but AHP not suspected* would benefit from AHP testing as they displayed suspicion of AHP or symptom complexes associated with AHP but have yet received a full diagnostic work-up.

Figure 1 shows a flowchart of the overall patient record filtering and manual review process. The process starts with 204,413 patient records, and using a combination of machine learning and structured data filtering described above, identifies 200 patients that were manually reviewed. 100 of those patients were identified as not having any mention of porphyria in the medical record and potentially could benefit from AHP diagnostic testing. The other 100 of those patients did have mention of porphyria in their medical record, but no diagnostic code for porphyria. These records were reviewed to determine the reason for the mention of porphyria and evaluate whether these reasons were consistent with the goal of the machine learning to identify patients with symptoms and other clinical features consistent with a possible porphyria diagnosis.

**Figure 1.**
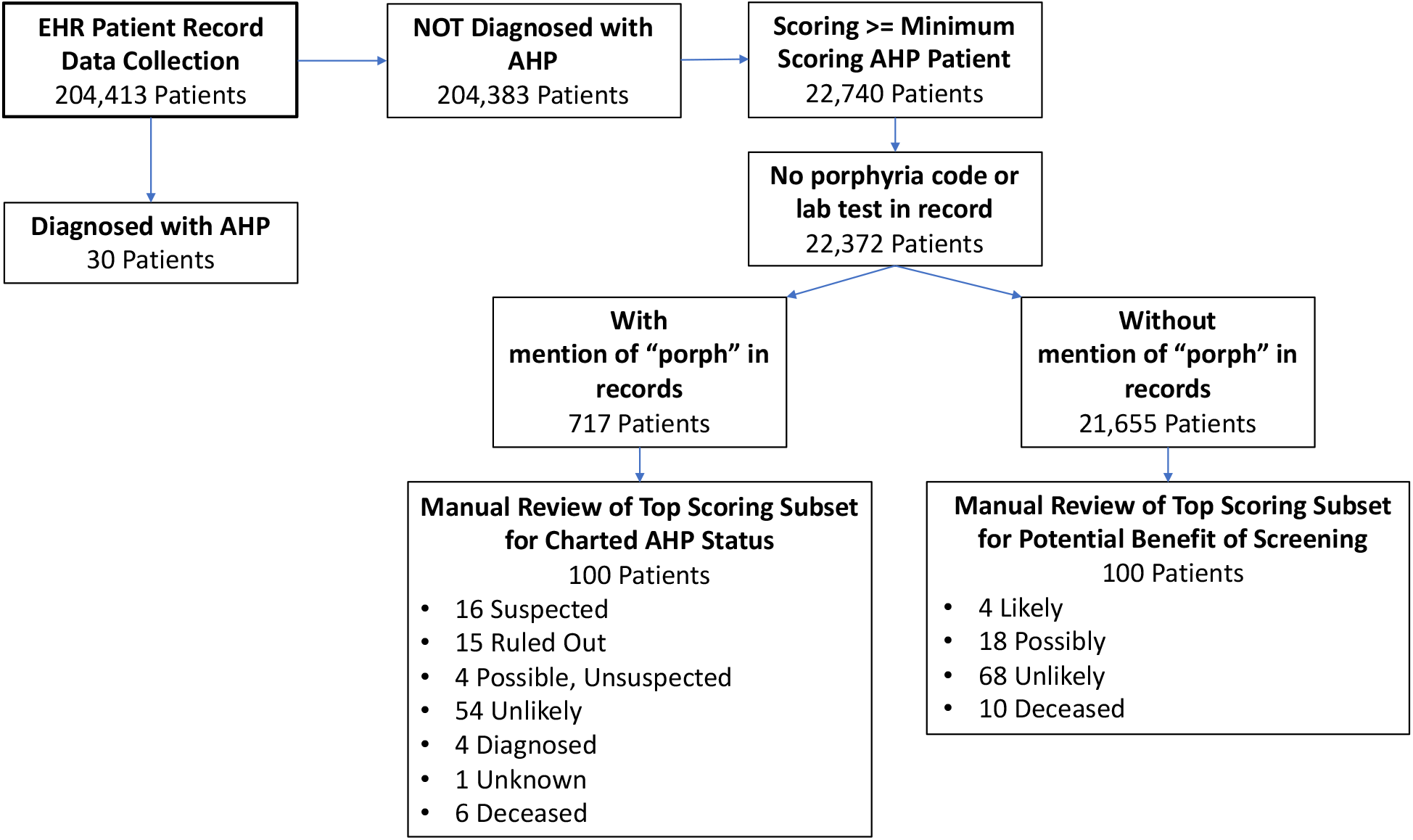
Flowchart of patient data record selection. Collection starts from full set of from full collection 204, 413 patient records and is filtered down to two sets of 100 records that were manually reviewed and characterized for 1) present indications for screening for AHP, and 2) status of AHP evaluation in the clinical notes of the record.

## Results

Out of the 100 patient charts we reviewed with no mention of porphyria, four were identified as likely to *AHP diagnostic testing likely indicated*, all without mention of porphyria in their medical record or documentation of a urine PBG test. The first patient was a male with six years of unexplained intermittent abdominal pain with nausea, vomiting, and diarrhea. His other conditions included complex regional pain syndrome, peripheral neuropathy, cardiac arrhythmias, panic attacks, and depression. The next patient was a female whose abdominal pain was described as ‘a long standing symptom with extensive negative evaluation’. Also listed in her profile were neuralgias, hereditary small fiber neuropathy, movement disorder, fibromyalgia, migraines, palpitations, and somatization disorder. The third patient was a woman with multiple emergency department admissions for severe abdominal pain. She also had severe suicidality with a permanent tracheostomy due to a hanging attempt, borderline personality disorder, tachycardia, anxiety, saddle anesthesia, insomnia, and severe somatization disorder including a comment in her note advising not to admit the patient for only vague complaints. The fourth patient was a female with a history of abdominal pain comments in the notes describing that the etiology had not been identified for her complex symptomology which included headaches, abdominal pain, paresthesias and palpitations.

Overall, about a quarter of the 100 patients in the group without mention of porphyria had symptom profiles that were consistent with undiagnosed AHP and AHP diagnostic testing would either be likely or possibly indicated (**Table 5**). In this group there was no sign or suspicion of AHP by the clinician in the record. This is a much higher concentration of possible AHP patients than would be expected by chance based on the known prevlance of AHP.

**Table 5.**
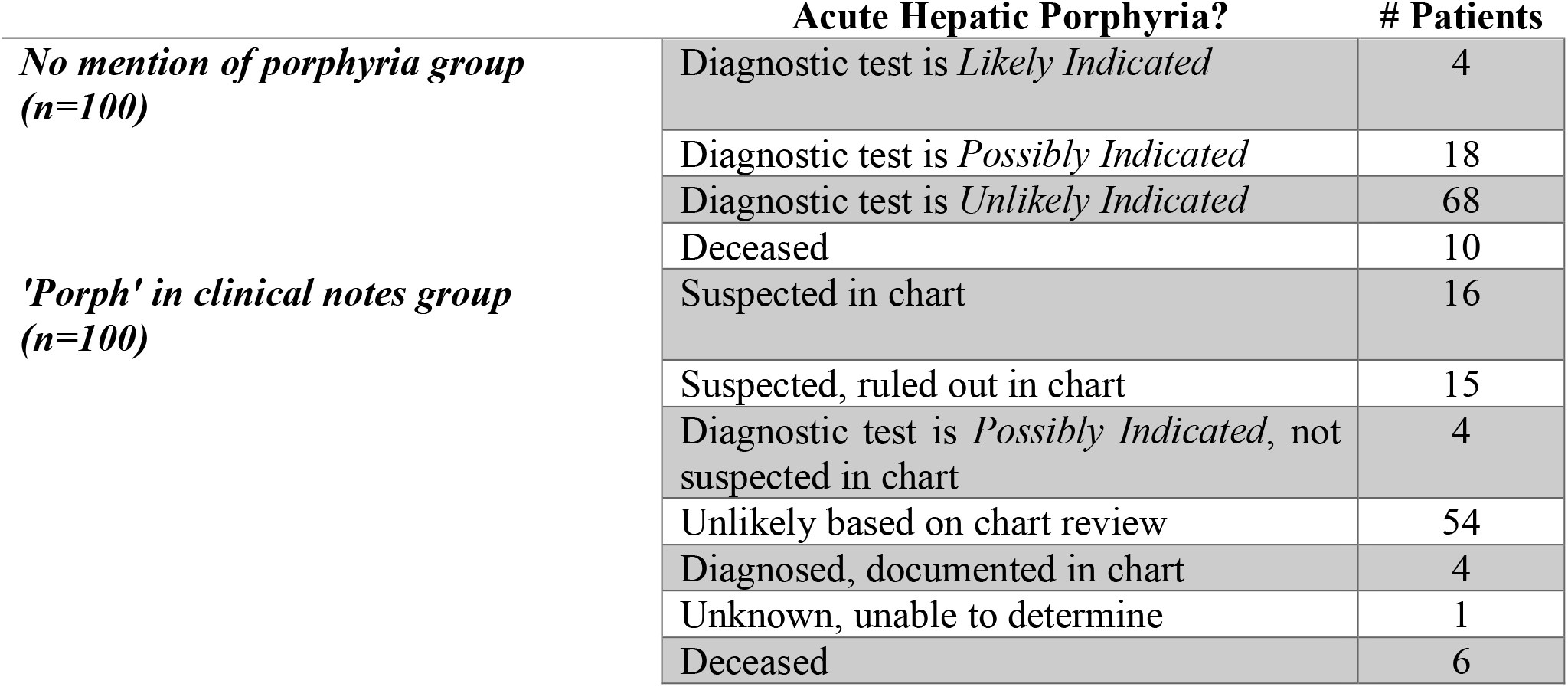
Assessment of the likelihood of undiagnosed acute hepatic porphyria based on clinical note symptom documentation. Both groups of 100 reviewed patients are listed.

Alternate explanations for characteristic AHP symptom profiles were diverse in the patient group without any mention of porphyria (**Table 6)**. Cancers seen in this group included breast, uterine, pancreatic, cervical, leukemia and adrenal carcinoma. Other common comorbidities and conditions seen in this group included: fibromyalgia, irritable bowel syndrome, chronic fatigue, obesity, hypertension, obstructive sleep apnea, and chronic obstructive pulmonary disease. In contrast, alternate symptom profiles in the group with mention of porphyria in the notes were dominated by liver pathologies, mostly hepatocellular carcinoma.

**Table 6.** Top alternative explanations for AHP symptom profiles seen in both groups of patients. Conditions seen in no more than one patient are not listed.

|  | Alternate AHP Symptom Explanation | # Patients |
| --- | --- | --- |
| *No mention of porphyria group* | Surgery | 8 |
|  | Inflammatory Bowel Disease | 6 |
|  | Cancer | 6 |
|  | Cancer Chemotherapy | 5 |
|  | Gallbladder Pathology | 4 |
|  | Diabetes | 3 |
|  | Carnitine Palmitoyl Transferase Deficiency | 2 |
|  | Renal | 4 |
|  | Poly Cystic Ovarian Syndrome | 2 |
|  | Appendicitis | 2 |
|  | Mastocytosis | 2 |
| *'Porph' in clinical notes group* | Liver Pathology | 30 |
|  | Chemotherapy/Drug Side Effects | 3 |
|  | Mastocytosis | 2 |

Patients in the group *without* mention of porphyria in the medical record generally had much longer and more complicated histories compared to the other group, with 86 out of 100 having encounters spread over four years or longer. The patients *with* porphyria mentioned in the clinical notes tended to have shorter, and less complex histories (only 39 out of 100 had over 4 years of encounters), more focused on a single medical issue or set of symptoms, which may have been due to their being referral to our academic medical center from other health care sites.

There were small differences in age summary statistics between the two groups (**Table 7**), but notably more pediatric patients in the reviewed group with mention of porphyria found in clinical notes than those without (10 patients vs 1 patient). There were significantly more male patients found in this group too, compared to the group with no mention of porphyria (**Table 8**). Associated conditions for these 44 male patients were dominated by only a few diagnoses/symptom patterns: liver disease (N=18), suspicion of porphyria (N=11), or actinic keratosis (N=3). In contrast, no single condition dominated the male disease distribution in the patient group without mention of porphyria in the notes.

**Table 7.** Age statistics in years for the two patient groups.

|  | **NO MENTION OF**<br>**PORPHYRIA** | **'PORPH' IN CLINICAL**<br>**NOTES** |
| --- | --- | --- |
| **MEDIAN** | 51 | 54 |
| **MEAN** | 53 | 50 |
| **MIN** | 8 | 6 |
| **MAX** | 91 | 91 |

**Table 8.** Sex distribution for the two patient groups.

|  | **NO MENTION OF**<br>**PORPHYRIA** | **'POPRH' IN CLINICAL**<br>**NOTES** |
| --- | --- | --- |
| **MALE** | 25 | 44 |
| **FEMALE** | 75 | 56 |

About a third of patients in the group *with* mention of porphyria in the clinical notes had some level of suspicion and work-up for AHP documented. We also identified four patients in this group that we thought had possibly undiagnosed AHP, without suspicion documented in the notes. We labeled these patients as *Diagnostic testing likely indicated but AHP not suspected*. Three of these patients had ‘porphyria’ in their clinical note listed as a standard precaution for several different medications (hydrochloroquinone, ferrous sulfate), which they were taking. In fact, about two thirds of the patients with ‘porphyria’ in the clinic notes had other reasons, besides suspicion of AHP, for the presence of this word (**Table 9**). A large number of these patients were candidates for liver transplantation. Standard clinical documentation for evaluation for this procedure included a list of possible causes of liver failure, including protoporphyria. Porphyria was also mentioned as a precaution for certain medications or treatments given to some patients in this group, which included hydroxycholorquinone ferrous sulfate, therapeutic abortion, and UV light therapy for actinic keratosis.

**Table 9.** Top reasons for the presence of the word ‘porph’ found in the clinical note.

| ***More Common Reasons for 'Porph' in Clinical Notes*** | **# Patients** |
| --- | --- |
| *Suspicion of Porphyria* | 31 |
| *Liver Transplant Documentation* | 30 |
| *Porphyria Mentioned in Treatment Precautions* | 18 |
| *Porphyria Diagnosis Mentioned in Notes* | 4 |
| *Porphyria Lab Tests Listed for Screening Physical* | 3 |
| *Family History of Porphyria* | 5 |
| *Misspelling* | 2 |

## Discussion

This work identified four likely and 18 possible patients who had no mention of porphyria in their charts for whom AHP diagnostic testing could be indicated. In addition, four patients who had mention of porphyria in their charts not related to a diagnostic evaluation of the disease were also found likely to have AHP diagnostic testing indicated. This number of patients with indications for AHP diagnostic testing and possibly to-be confirmed diagnosis vastly exceeds that due to chance and surpassed our expectations. It will require clinical follow-up to determine whether these patients’ symptoms are truly due to AHP or not, but the manual record review clearly demonstrates that our methodology has found patients for whom a spot urine porphobilinogen test is indicated.

Another benefit of identifying such patients is to inform local specialists of the presence of patients with rare diseases in which they have expertise. An institution-wide search for confirmed AHP patients through our targeted ICD-10-CM code search plus manual chart review identified 30 confirmed AHP patients. A majority of these patients were previously unknown to the porphyria specialist (TD) at OHSU. Identifying rare disease patients through large-scale data review in this manner can help connect them with the appropriate specialist to ensure optimal care.

Our results strongly suggest that leveraging of EHR data coupled with machine learning can be an effective method of identifying patients who should receive a diagnostic biochemical test to screen for AHP. Our automated model was able to identify patients with compelling constellations of symptoms who had not be previously worked up for porphyria. It was also able to identify patients for whom porphyria had been considered without direct access to porphyria-related data elements such as hemin treatment, lab tests specific to AHP, or mention of AHP diagnosis in clinical notes.

This is especially interesting in the light that the overall cross-validation scores of the model on the data set using the known 30 AHP cases as the positive set and the rest of the data as negative training samples was not very high, with cross-validation yielding an average AUC = 0.775. This is certainly a low performance figure compared to other current machine learning tasks such as publication type identification [24], or facial image recognition [25]. However, these other tasks are very different from this one due to the extremely rare nature of the positive AIP cases in both the training data as well as in the actual patient population. In most machine learning research, a data set is considered skewed or imbalanced if the number of positive cases is much less than 50%. A recent systematic review on imbalanced data classification cites articles investigating negative to positive case ratios of 100 to 1 as “highly imbalanced” [26, 27]. For problems such as rare diseases, the imbalance ratio can be nearly 10,000 to 1, as it is here. Lifting the predictive power to perhaps 22 in 100 manually reviewed cases is a potentially transformative level of performance.

The strongest positive predictors in the model included unexplained abdominal pain, pelvic and perineal pain, nausea and vomiting, and a number of pain and nausea medications. Frequent urinalysis was also a strong positive predictive feature, this is likely due to being associated with frequent ER visits and hospitalizations. The model relied on encoding the frequency of episodes, and not just binary presence of absence of symptoms. Indirectly, in the model this represented recurrent, undiagnosed problems consistent with AHP.

As these methods are general, and not specific to AHP, they should be applicable to other rare disorders that have a constellation of recurrent symptoms as indicating features. There are likely ways to improve the machine learning approach, including the use of more advanced features that represent time, duration, and intervals, explicit coding of symptom separation and overlap, and more sophisticated machine learning algorithms specifically tailored to situations where the positive case is extremely rare. Investigation into machine learning algorithms for highly skewed data such as these is an active area of research [28].

## Conclusion

The combination of large data sets, machine learning techniques, and clinical knowledge engineering can be a powerful tool to identify patients with undiagnosed rare diseases. The use case of AHP presented here revealed four undiagnosed patients thought likely to have AHP, as well as 18 others who would likely benefit from testing. This level of precision in identifying potential cases of AHP from EHR data is much higher than would be expected by the prevalence of the disease.

Analyzing the EHR with advanced techniques such as demonstrated here points to the potential of the future of digital medicine on a population scale. Advanced approaches enabled by the wide deployment of the EHR can now be used to improve medicine and medical care in areas that have been underserved or inaccessible. Health care can be made more proactive, not simply in terms of common conditions and age or gender related screening, but for rarer conditions as well.

We plan to continue this work in several directions. First, an IRB-approved clinical validation study is being implemented. In this study, we will contact the primary care clinicians (PCP) of the patients where AHP diagnostic testing was found to be *likely* or *possibly* indicated. We will inform them that an algorithm based on EHR data has determined that their patient might have AHP and could benefit from a spot urine porphobilinogen, which is an is inexpensive, non-invasive and easy to perform diagnostic test. With the agreement of the PCP, we will then contact patients and offer them the test. Expert clinical consultation will be made available to the PCP for any questions they have. We will collect data on the interactions with the PCPs, the number of spot urine porphobilinogen tests administered, as well as the test results. In this manner, we will be able to study the clinical impact of our rare disease identification approach. Second, we will continue to refine our methods. Other machine learning algorithms, such as random forests and deep learning, may have advantages for AHP and other rare diseases. Other methods of encoding the EHR data that incorporate embeddings and temporal representations, have been shown to demonstrate leading-edge results in other fields, such as computer vision, machine translation, and speech recognition, and may assist with rare diseases.

Finally, we will extend this methodology to other rare diseases that are difficult to diagnose, focusing on those for which effective treatments are becoming available. If the timeline for diagnosing rate conditions can be substantially reduced, there is great potential to impact patient health in a very significant manner.

## Data Availability

The electronic health record data used in this work is protected by HIPAA requirements, and cannot be made publically available.

## Acknowledgements and Funding

This work was funded and the associated editorial support was provided by Alnylam Pharmaceuticals, Inc., Cambridge, MA.

## Declaration of Interest

Stephen Meninger, John J. Ko, and Jigar Amin, are employees of Alnylam, and Alex Wei was an employee of Alnylam during his contribution to the manuscript.

**Supplemental Table 1.**
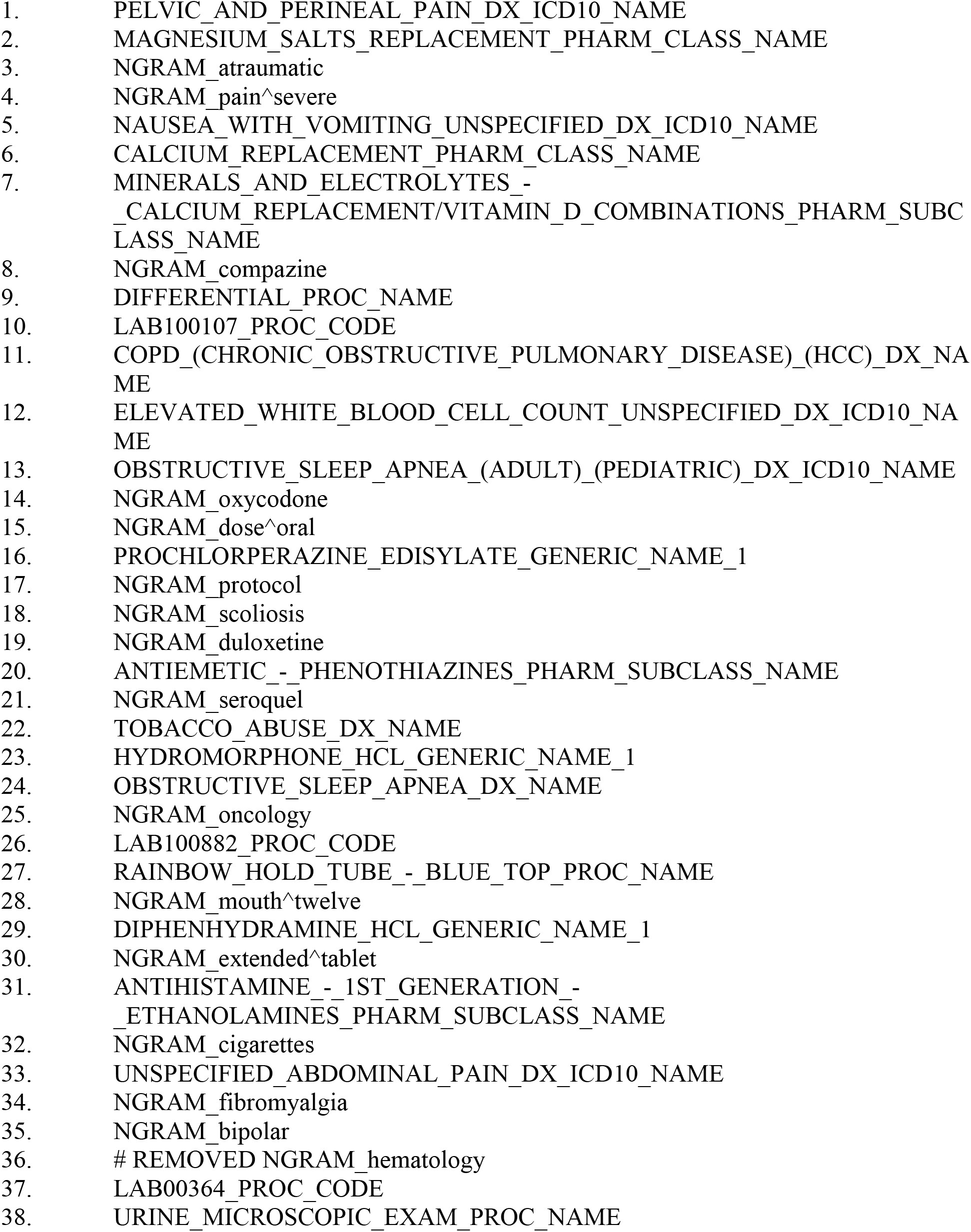
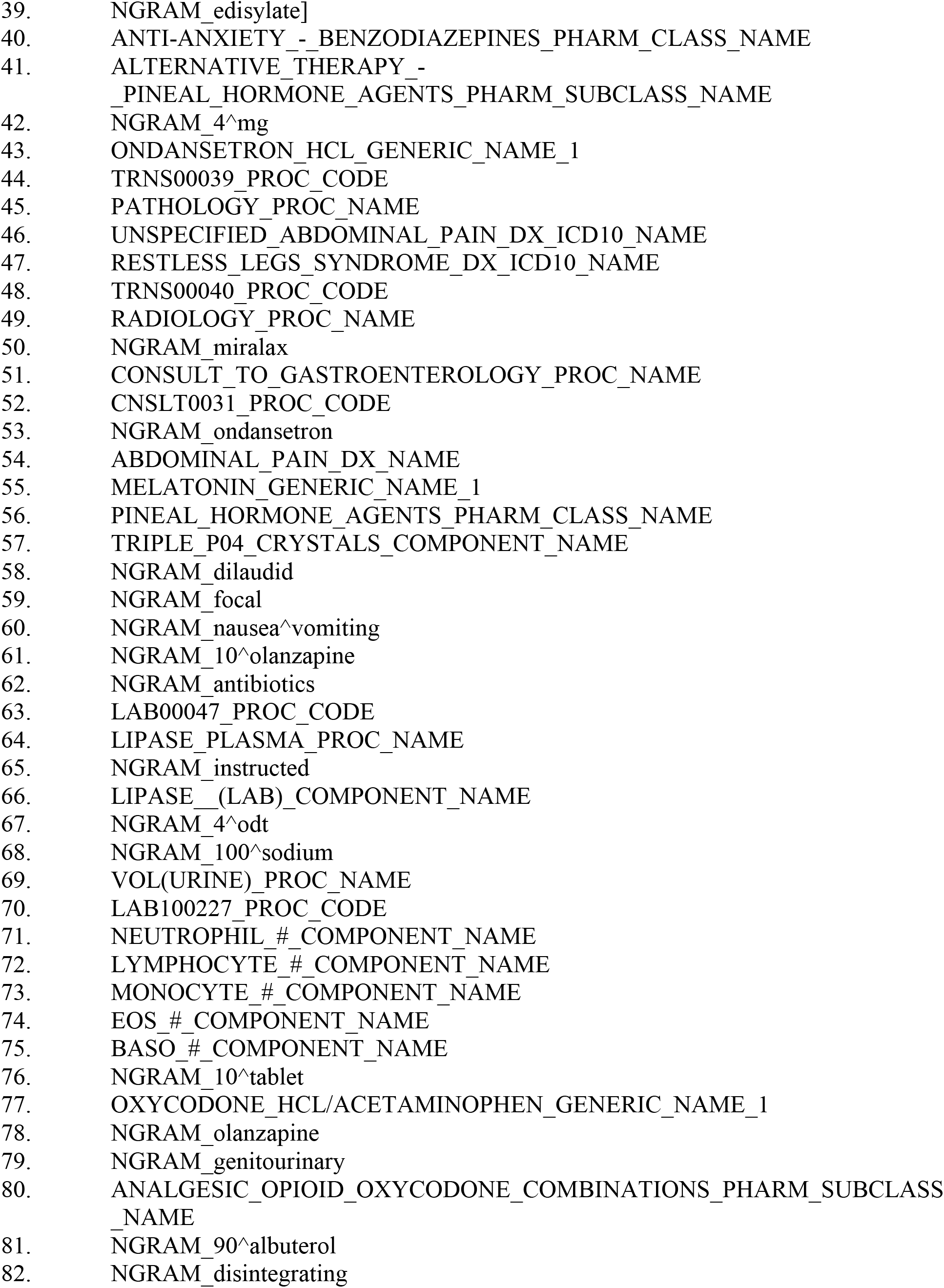
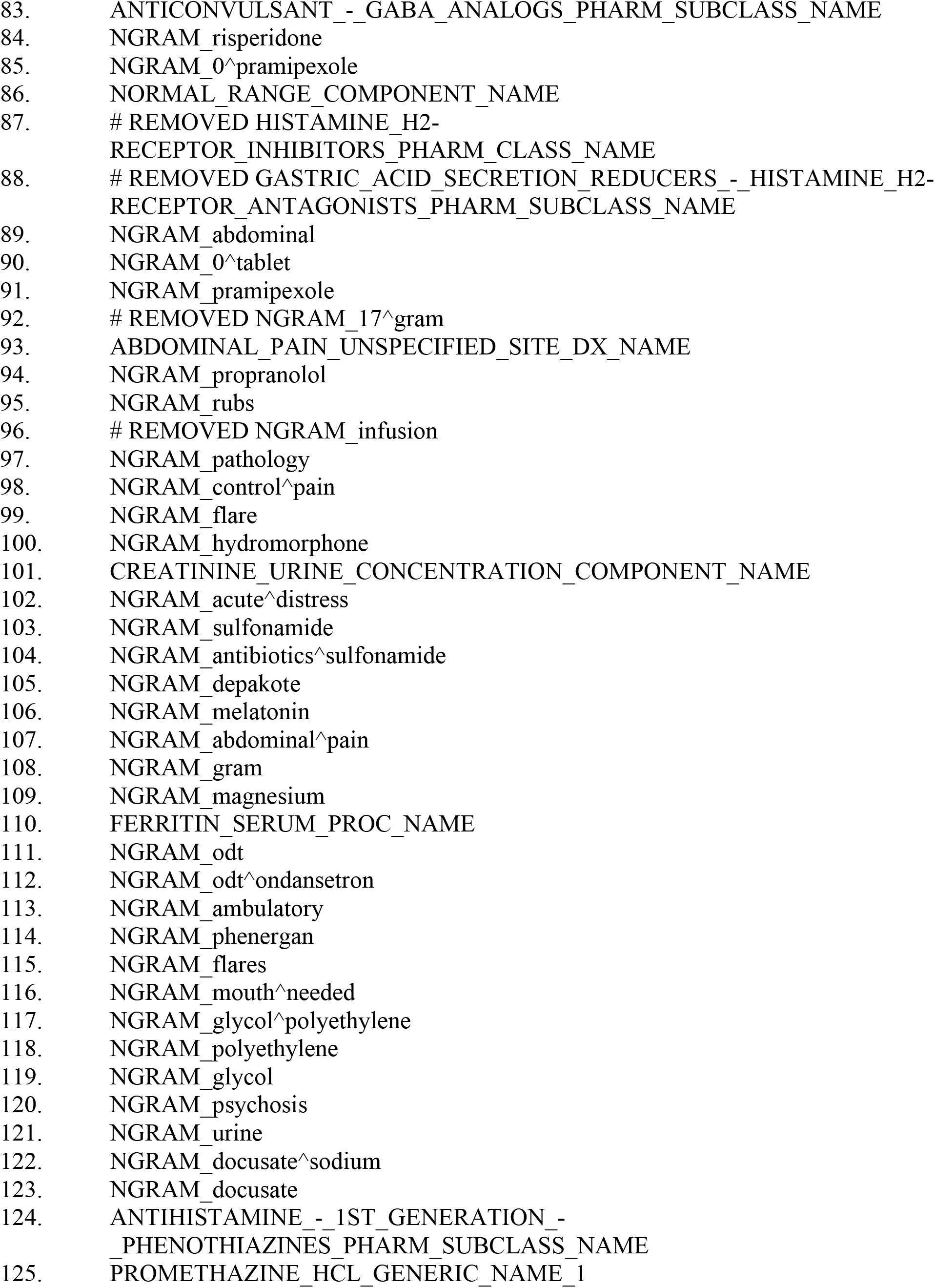
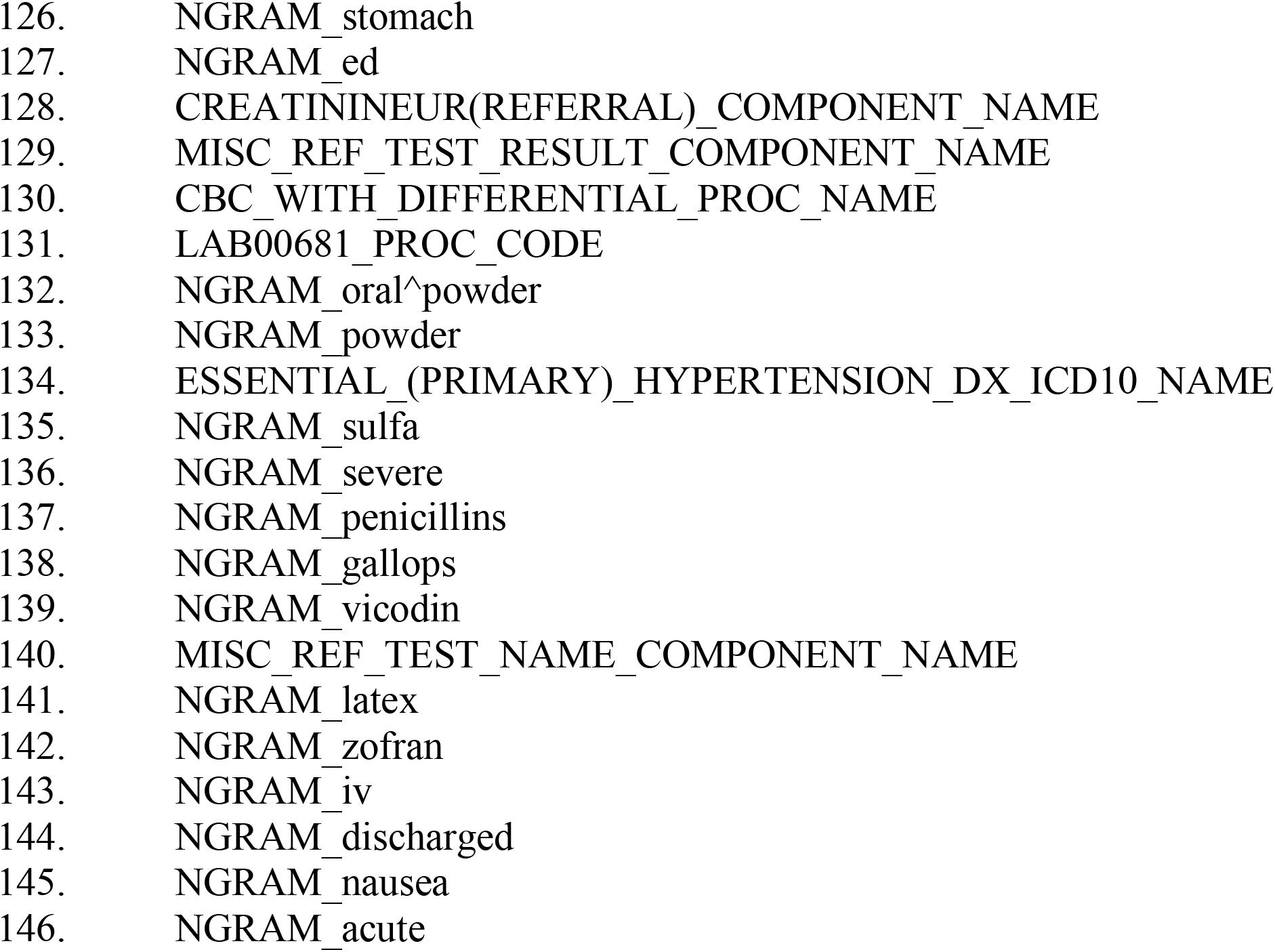
Final 146 features selected for inclusion in the machine learning model to predict acute hepatic porphyria.

